# Evaluation of primary allied healthcare in patients recovering from COVID-19: first results after six months follow-up in a Dutch nationwide prospective cohort study

**DOI:** 10.1101/2022.10.03.22280639

**Authors:** Anne I. Slotegraaf, Marissa H.G. Gerards, Arie C. Verburg, Marian A.E. de van der Schueren, Hinke M. Kruizenga, Maud J.L. Graff, Edith H.C. Cup, Johanna G. Kalf, Antoine F. Lenssen, Willemijn M. Meijer, Renée A. Kool, Rob A. de Bie, Philip J. van der Wees, Thomas J. Hoogeboom, the Dutch Consortium Allied Healthcare COVID-19

## Abstract

**Objectives:** To report the recovery of patients receiving primary allied healthcare after a COVID-19 infection at a six-month follow-up, and to explore which patient characteristics are associated with the changes in outcomes between the baseline and six-month follow-up.

**Design:** Prospective cohort study.

**Setting:** Allied healthcare in Dutch primary care.

**Participants:** 1,451 adult patients recovering from COVID-19 and receiving treatment from one or more primary care allied health professional(s) (i.e., dietitian, exercise therapist, occupational therapist, physical therapist and/or speech and language therapist).

**Results:** For participation (USER-P range 0 to 100), estimated mean differences of at least 2.3 points were observed after six months. For HRQoL (EQ-VAS range 0 to 100), the mean increase was 12.31 at six months. Furthermore, significant improvements were found for fatigue (FSS range 1 to 7): the mean decrease was –0.7 at six months. For physical functioning (PROMIS-PF range 13.8 to 61.3), the mean increase was 5.9 at six months. Mean differences of –0.8 for anxiety (HADS range 0 to 21), and –1.5 for depression (HADS range 0 to 21), were found after six months. Having a worse baseline score, hospital admission and male sex were associated with greater improvement between the baseline and six-month follow-up, whereas age, BMI, comorbidities and smoking status were not associated with mean changes in any outcome measure.

**Conclusions:** Patients recovering from COVID-19 who receive primary allied healthcare make progress in recovery, but still experience many limitations in their daily activities after six months. Our findings provide reference values to healthcare providers and healthcare policy-makers regarding what to expect from the recovery of patients who received health care from one or more primary care allied health professionals.

**Trial registration:** Clinicaltrials.gov registry (NCT04735744).

## Introduction

An estimated 32–57% of patients recovering from a COVID-19 infection experience severe and long-term problems in daily functioning and participation.[1-3] It is becoming increasingly clear that both patients with mild symptoms and those with serious symptoms during the acute COVID-19 infection are at risk of developing post-COVID-19 syndrome.[1, 2, 4, 5] Post-COVID-19 syndrome, also referred to as ‘long COVID’, is defined as ‘signs and symptoms that develop during or after a COVID-19 infection, continuing for more than 12 weeks, [that] are not explained by an alternative diagnosis’.[6-8] To date, it is unknown what treatment is needed to support patients in their recovery from COVID-19.

Patients recovering from COVID-19 often experience persistent problems in their daily activities related to limitations in physical, nutritional, cognitive and mental functioning.[3, 5, 9-11] Fatigue is the most prevalent and persistent symptom, irrespective of the severity of the initial infection.[3, 5, 10, 12, 13] Longitudinal data suggest that fatigue does not resolve over time in many patients, even if they receive healthcare.[3, 9, 10, 13, 14] Increased levels of fatigue can result in lower levels of physical activity [15] and limit patients in activities of daily living (e.g., housekeeping and grocery shopping) and outdoor pursuits.[16] Mental problems such as anxiety and depression are common in patients recovering from COVID-19. A study by Huang et al. (2021) showed that anxiety, depression and sleep difficulties were present in approximately 25% of patients six months after the onset of COVID-19 symptoms.[17] Sisó-Almirall et al. (2021) showed that 36% of patients still reported mental problems after three months, and no significant associations were found with COVID-19 severity.[10] Furthermore, previous studies observed a worsened health-related quality of life (HRQoL) in patients recovering from COVID-19, both hospitalised and non-hospitalised, which did not recuperate after a follow-up period of several months.[9, 18-21]

Based on the overall effects of primary allied healthcare, it is expected that allied health professionals (i.e. dietitians, exercise therapists, occupational therapists, physical therapists and speech and language therapists) can play a role in the recovery of patients with COVID-19 who experience persistent limitations in daily physical functioning and participation.[22-25] In July 2020, the Dutch Ministry of Health, Welfare and Sports instated a temporary regulation in primary allied health care to facilitate the treatment of patients recovering from COVID-19 and to stimulate research. This regulation enables the reimbursement of primary allied health care for every patient from basic health insurance coverage. With a referral from a general practitioner (GP) or medical specialist, primary allied healthcare treatment is reimbursed for a period of six months. If recovery during this period is insufficient, an extension by a second six-month period is possible upon referral by a medical specialist. As COVID-19 is still a novel condition and the evidence base for allied health treatment in patients with post-COVID-19 syndrome is small, it is vital that new data and insights are shared as soon as they are available; therefore, the aim of the current paper is to present the results of recovery by patients receiving primary allied healthcare after a COVID-19 infection. We provide outcomes at three- and six-months follow-ups regarding participation, HRQoL, physical functioning, fatigue and psychological well-being. In addition, we explore which baseline characteristics are associated with changes in these outcomes between the baseline and six-month follow-up.

## Methods

### Study design and setting

As part of a nationwide project to evaluate the recovery of patients receiving primary allied healthcare after a COVID-19 infection, a prospective cohort study was set up in collaboration with various patient organisations (i.e., the Lung Foundation Netherlands, the Netherlands Patient Federation and Harteraad) and with input from patients contacted through these organisations.[26] In this prospective cohort study, patients were included at the start of their treatment with one or more allied health professionals. The patients received follow-ups at three-month intervals for one year after inclusion. All treatment trajectories offered by allied health professionals in daily practice were part of usual care, and were preferably based on recommendations and guidelines published by the professional bodies of the respective care providers as available at the start of the research.[22-25] The inclusion period for the cohort study was between 29^th^ March 2021 and 19^th^ June 2021. Primary outcome measures were assessed at the baseline (T0), and again after three months (T1), six months (T2), nine months (T3) and 12 months (T4). The primary endpoint of the study was set at six months (T2). In the present paper, we will report the results of our primary outcome measures at the three- and six-month follow-ups.

The study protocol was reviewed and approved by the medical ethics committee of Radboud university medical centre (Registration #2020-7278). The study has been registered in the clinicaltrials.gov registry (NCT04735744). Informed consent was obtained from all patients before enrolment in the study.

### Participants

Adult patients (aged ≥18 years) were eligible for inclusion in the cohort if they were recovering from COVID-19 and started treatment with one or more primary care allied health professionals (i.e., a dietitian, exercise therapist, occupational therapist, physical therapist and/or speech and language therapist). Patients may have followed trajectories with one or more allied health professionals during the course of the study. Patients were included regardless of their hospital admission status during the acute phase of COVID-19. Patients who were unable to complete questionnaires in Dutch and patients who were receiving palliative care were excluded from the study.

### Data collection

Patients could enrol in the study by a) signing up after an invitation by their treating allied health professional, or b) signing up on their own initiative, upon which the research team also invited the treating allied health professional to participate. The enrolment procedure of this study is described in detail in the published study protocol.[26] Both patients and allied health professionals reported data via the specifically designed YourResearch® application. Patients were asked to download an application on their smartphones or make use of a web application. Questionnaires were sent out through this application at the start of the treatment (baseline), and after three, six, nine and 12 months. Patients unable to participate via digital methods were given the opportunity to complete the questionnaires on paper and return them by post. Allied health professionals were asked to use a web application and report data on patient characteristics and profession-specific outcome measures. These profession-specific outcome measures are not included in the present paper but will be reported elsewhere.

### Outcome measures

Data on patient characteristics were collected by the treating allied health professional at the start of the treatment. Patient-reported outcome domains participation, HRQoL, fatigue and physical functioning were assessed at the baseline, and after three and six months. Data on psychological well-being were collected at the baseline and after six months.

#### Patient characteristics

Patient characteristics were collected via an online record form and contained the following items on demographics: age, sex, height (in cm), weight (in kg) both at the start of treatment and before COVID-19 infection, living status and whether the patient had an informal caregiver. Furthermore, data on symptom severity at the onset of treatment (i.e., mild to moderate (mild symptoms up to mild pneumonia), severe (dyspnoea, hypoxia, or <50% lung involvement on imaging), or critical (respiratory failure, shock, or multiorgan system dysfunction), definitions as described in [27]), as well as hospital admissions during the acute phase of COVID-19 (i.e., no hospital admission, admission to hospital ward or intensive care unit), were recorded. The following items were included about the allied healthcare treatment trajectory: referring physician, treatment goals, and mono- or multidisciplinary treatment. Additionally, data on comorbidities (i.e., cardiovascular disease, chronic lung disease, diabetes mellitus, kidney disease, liver disease, immune disease, oncological disease, chronic neuromuscular disorders) and smoking status were collected. Body weight and height were used to calculate each patient’s body mass index (BMI) (weight/height^2^) and was categorised as defined by the World Health Organisation (WHO).[28]

#### Participation

Participation was assessed with the Utrecht Scale for Evaluation of Rehabilitation-Participation (USER-P). The USER-P is a 31-item self-administered questionnaire reflecting a patient’s participation in daily life, divided over three subscales: frequencies, restrictions and satisfaction. The total scores range from 0 to 100 for each subscale, with higher scores indicating better participation (higher frequency, fewer restrictions and higher satisfaction).[29] We arbitrarily assumed a 5.0-point difference on one of these USER-P scales to be clinically relevant for patients recovering from COVID-19.

#### Health-related quality of life

HRQoL was assessed with the EQ-5D-5L, a five-item questionnaire measuring a person’s status in five dimensions of health: mobility, self-care, usual activities, pain/discomfort and anxiety/depression.[30] Furthermore, the EQ visual analogue scale (VAS) was recorded by the patients. The EQ-VAS provides a quantitative measure of the patient’s perception of their overall health in a score ranging from 0 to 100, with higher scores indicating a higher HRQoL. A difference of 8.0 points on the EQ-VAS was considered clinically relevant.[31]

#### Fatigue

Fatigue was assessed with the Fatigue Severity Scale (FSS), a nine-item scale measuring the severity of fatigue and its effect on patients’ activities and lifestyle. the scoring of each item ranges from 1 to 7, where 1 indicates strong disagreement and 7 indicates strong agreement. The total score is calculated by the mean value of the nine items, with a score of 4 or more indicating severe fatigue.[32] A difference of 0.45 points on the FSS mean score was considered clinically relevant.[33]

#### Physical functioning

Limitations in physical functioning were assessed with the PROMIS Physical Functioning Short Form 10b (PROMIS-PF-10b), a 10-item questionnaire measuring the self-reported ability to perform the activities of daily life. Items reflect four subcategories: upper extremities (dexterity), lower extremities (walking or mobility), and central regions (neck and back), as well as instrumental activities of daily living, such as running errands.[34] Total scores range from 13.8 (severely physically impaired) to 61.3 (not physically impaired), with a mean score of 50 (SD 10) representing the mean score of a reference population.[35] A difference of 3.6 points was considered clinically relevant.[36]

#### Psychological well-being

The Hospital Anxiety and Depression Scale (HADS) was used to assess psychological well-being. This 14-item self-administered questionnaire describes symptoms of anxiety and depression. The HADS is divided into an Anxiety score (HADS-A) and a Depression score (HADS-D), each containing seven items. The total score ranges from 0 to 21 for both subscales, where a total score of 11 or more indicates a probable clinical diagnosis of depression or anxiety.[37, 38] A difference of 1.7 points was considered clinically relevant.[39]

### Statistical analyses

Descriptive statistics were used to describe the patient population and to analyse the primary outcome measures at the baseline and after three and six months, using numbers and proportions for categorical variables, means with standard deviations (SD), and medians with interquartile ranges (IQR) for continuous variables. Linear mixed model analyses were used to evaluate recovery over time for participation, HRQoL, fatigue and physical functioning. This analysis accounts for correlation between repeated measures on the same subject and uses all available data from this subject. A model with a random intercept and all other variables fixed was also generated. The primary outcomes were used as dependent variables, while time (categorical: baseline, three and six months) was used as a fixed factor. Paired samples t-tests were performed to evaluate recovery of psychological well-being at the six-month follow-up.

Uni- and multivariate regression analyses were used to explore which baseline characteristics were associated with the change in score for the main outcome measures between the baseline (T0) and the six-month follow-up (T2). This analysis used data from complete cases, and missingness at random (MAR) was tested (Supplemental File 1). Univariate analyses were performed to determine which baseline characteristics (i.e., age, sex, BMI, hospitalisation, comorbidities, baseline score and smoking status) were associated with the mean change in each outcome measure. Comorbidities were coded into three categories: none, 1, and 2 or more comorbidities. Variables with *p* < 0.157 in the univariate regression were included in the multivariate model [40]. The backward elimination of variables was then performed in order of statistical significance until only factors that were significantly associated with the outcome remained. A p-value <0.05 was considered statistically significant for all analyses, based on two-sided testing. All data were analysed using SPSS statistics 25(IBM).

### Patient and public involvement

During the development of the current study, we involved patients to give feedback on the readability and appropriateness of proposed measures. The usability of the smartphone and web-based applications was also tested by patients. Participating patients received updates on the status of the study via their smartphone or the web application. Furthermore, various patient organisations (i.e., the Lung Foundation Netherlands, the Netherlands Patient Federation and Harteraad) participated during routine research meetings.

## Results

### Patient characteristics

In total, 1,451 patients were included in this study (Figure 1), receiving 1,708 different allied healthcare treatment trajectories. Their mean age was 49 (SD 13) years, and 64% of the patient population was female (Table 1). The majority (77%) had not been hospitalised for COVID-19, and 76% had experienced a mild to moderate severity of symptoms during the infection period. The mean (SD) BMI was 28 (6) kg/m^2^, and 69% of the patient population was classified as being overweight or obese (BMI > 25 kg/m^2^). One comorbidity was reported by 31% of the patients, and two or more comorbidities were reported by 12% of the patients. Cardiovascular disease (15%) and chronic lung disease (14%) were the most prevalent comorbidities. Most patients (82%) had been referred for primary allied healthcare by their GP.

**Table 1.** General characteristics of patients recovering from COVID-19 receiving primary allied healthcare.

|  | **Total group**<br>(n = 1451) |
| --- | --- |
| **Treatment trajectories, n (%)** | 1708 |
| Physical therapy/exercise therapy | 1005 (59) |
| Occupational therapy | 364 (21) |
| Dietary care | 224 (13) |
| Speech and language therapy | 115 (7) |
| **Sex, n (%)** | <sup>a</sup>n = 1330 |
| Male | 482 (36) |
| Female | 848 (64) |
| **Age, mean ± SD** | <sup>a</sup>n = 1331<br>49 ± 13 |
| **COVID-19 severity, n (%)** | <sup>a</sup>n = 1311 |
| Mild/moderate | 1002 (76) |
| Serious | 271 (21) |
| Very serious | 38 (3) |
| **Admission to hospital for COVID-19 infection, n (%)** | <sup>a</sup>n = 1315 |
| Hospitalised including intensive care | 87 (7) |
| Hospitalised | 213 (16) |
| Not hospitalised | 1015 (77) |
| **BMI, mean ± SD** | <sup>a</sup>n = 1071<br>28 ± 6 |
| Underweight (<18.5 kg/m<sup>2</sup>) | 10 (1) |
| Normal weight (18.5–25 kg/m<sup>2</sup>) | 323 (30) |
| Overweight (25–30 kg/m<sup>2</sup>) | 404 (38) |
| Obesity (>30 kg/m<sup>2</sup>) | 334 (31) |
| **Smoking status, n (%)** | <sup>a</sup>n = 1305 |
| Current | 63 (5) |
| Former | 166 (13) |
| Never | 1076 (82) |
| **Living status, n (%)** | <sup>a</sup>n = 1322 |
| Alone | 212 (16) |
| Cohabiting | 1110 (84) |
| **Informal carer, n (%)** | <sup>a</sup>n = 1319 |
| Yes | 526 (40) |
| No | 793 (60) |
| **Comorbidities, n (%)** | <sup>a</sup>n = 1331 |
| No comorbidities | 766 (57) |
| One comorbidity | 410 (31) |
| Two or more comorbidities | 155 (12) |

<sup>a</sup>Data were not fully available for all patients: the n within the table denotes the number of patients with available data.

**Figure 1.**
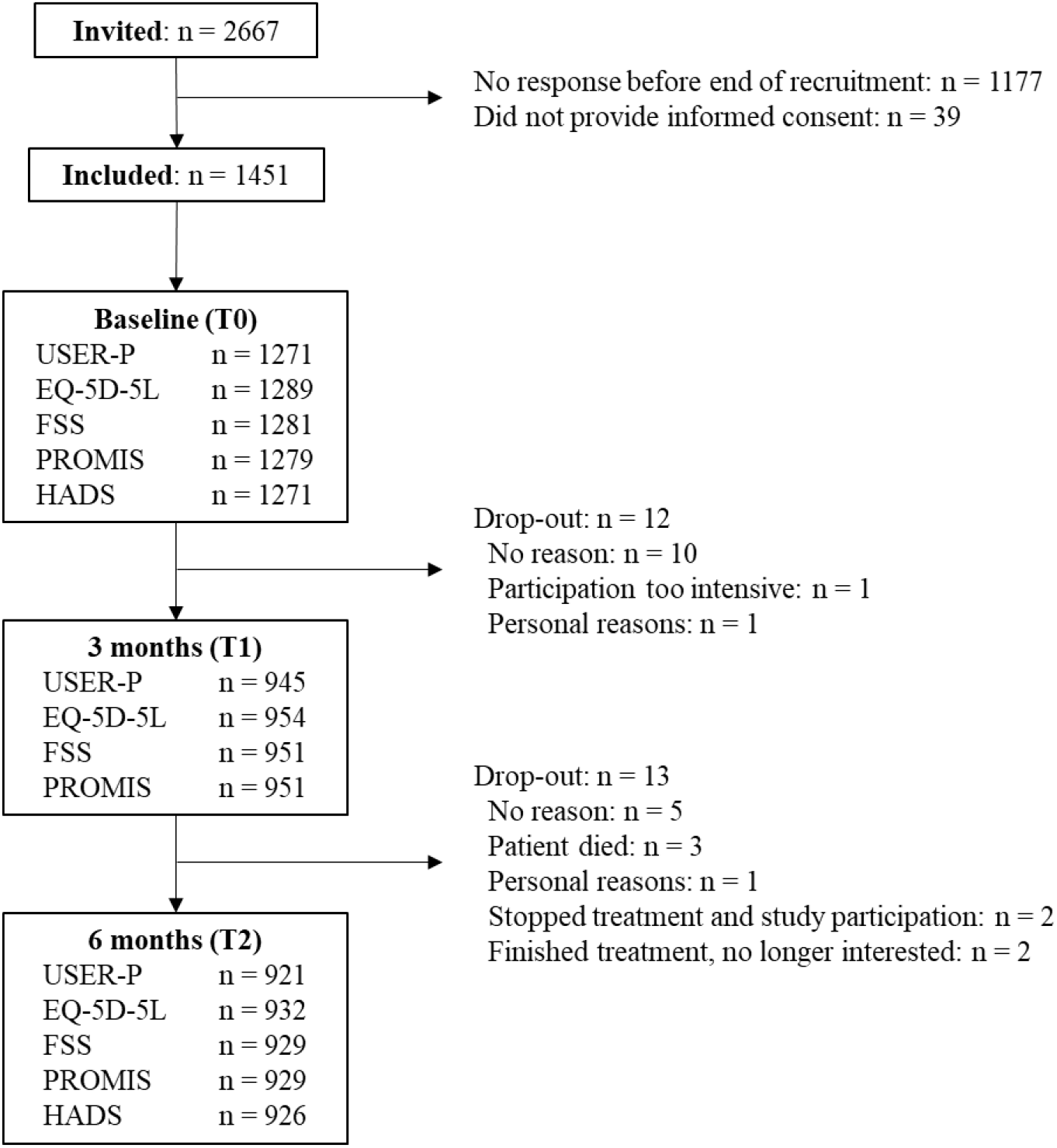
Flow diagram inclusion of patients recovering from COVID-19 receiving primary allied healthcare, with follow-ups after three and six months from the start of treatment. *Abbreviations:* USER-P: Utrecht Scale for Evaluation of Rehabilitation Participation. EQ-VAS: EuroQol Visual Analogue Scale. FSS: Fatigue Severity Scale. PROMIS: Patient-Reported Outcomes Measurement Information System. HADS: Hospital Anxiety and Depression Scale.

### Primary outcome measures

Table 2 presents data on the outcome measures at the baseline, and at the three- and six-month follow-ups. Additionally, clinically relevant improvements at the six-month follow-up are presented in Supplemental File 2. After six months, the majority of patients showed a clinically relevant improvement on the USER-P restrictions and satisfaction scales (65% and 61% of patients, respectively), while 60% of patients showed a clinically relevant improvement on the EQ-VAS (mean 67.4 (SD 19.1) points) compared with the baseline (55.5 (SD 17.8) points). Severe fatigue was reported by 94% of patients at the baseline, persisting after six months in 80% of patients. A clinically relevant improvement on the FSS mean score was found in 54% of patients. Based on PROMIS-PF-10b scores, over two thirds of the patients reported to be more than 60% impaired, limited or restricted in physical functioning at the baseline, which decreased to 38% after six months; 57% of patients experienced a clinically relevant improvement in physical functioning. The majority of patients scored less than 7 points on the HADS anxiety and depression scores both at the baseline and at six months, which indicates no anxiety disorder or depression. At the baseline, the HADS anxiety score indicated a probable clinical diagnosis of anxiety disorder in 23% of the patients, which decreased slightly to 18% after six months. A probable clinical diagnosis of depression was indicated by the HADS depression score in 22% of the patients at the baseline, decreasing to 15% at the six-month follow-up.

**Table 2.**
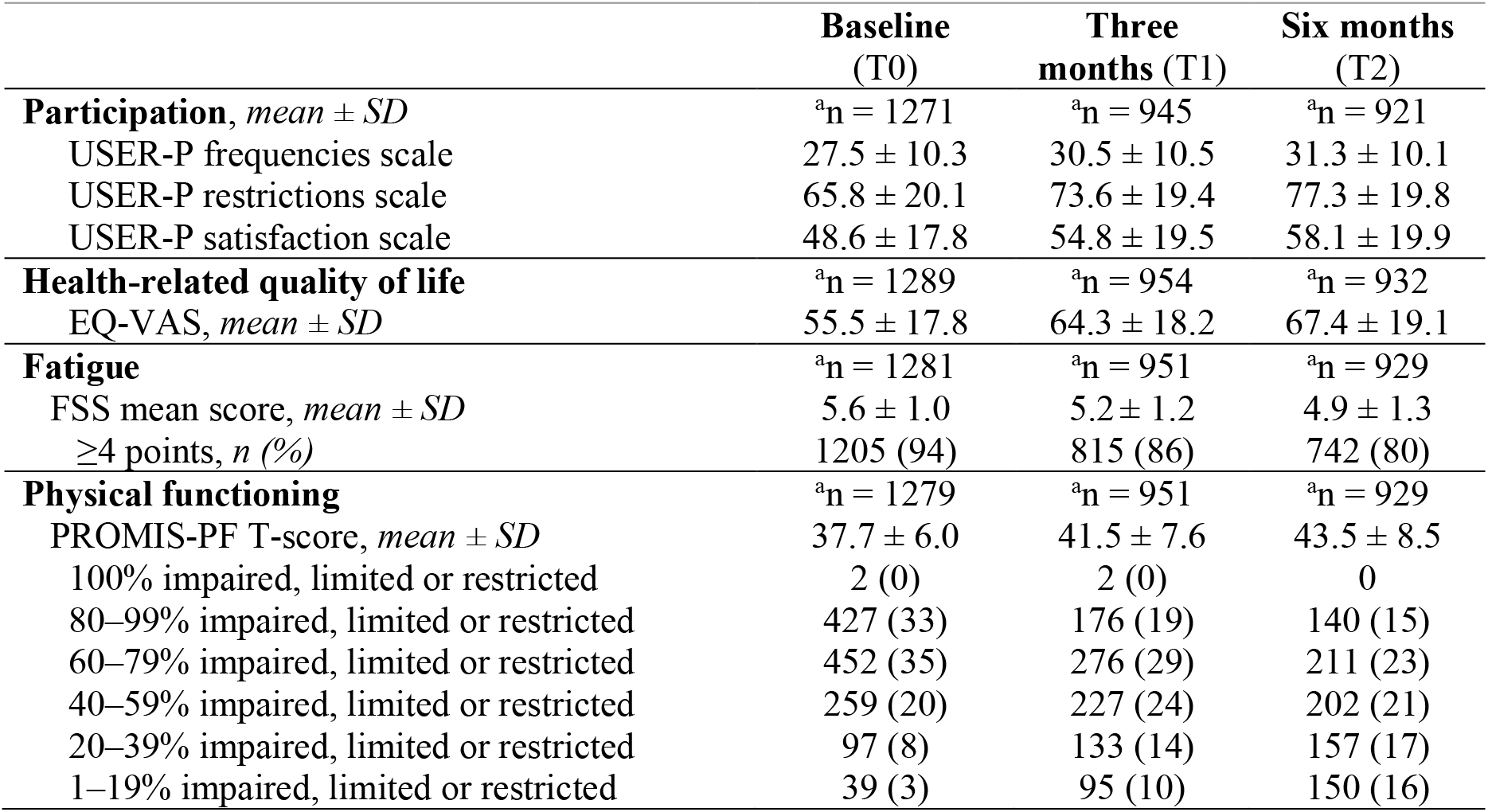
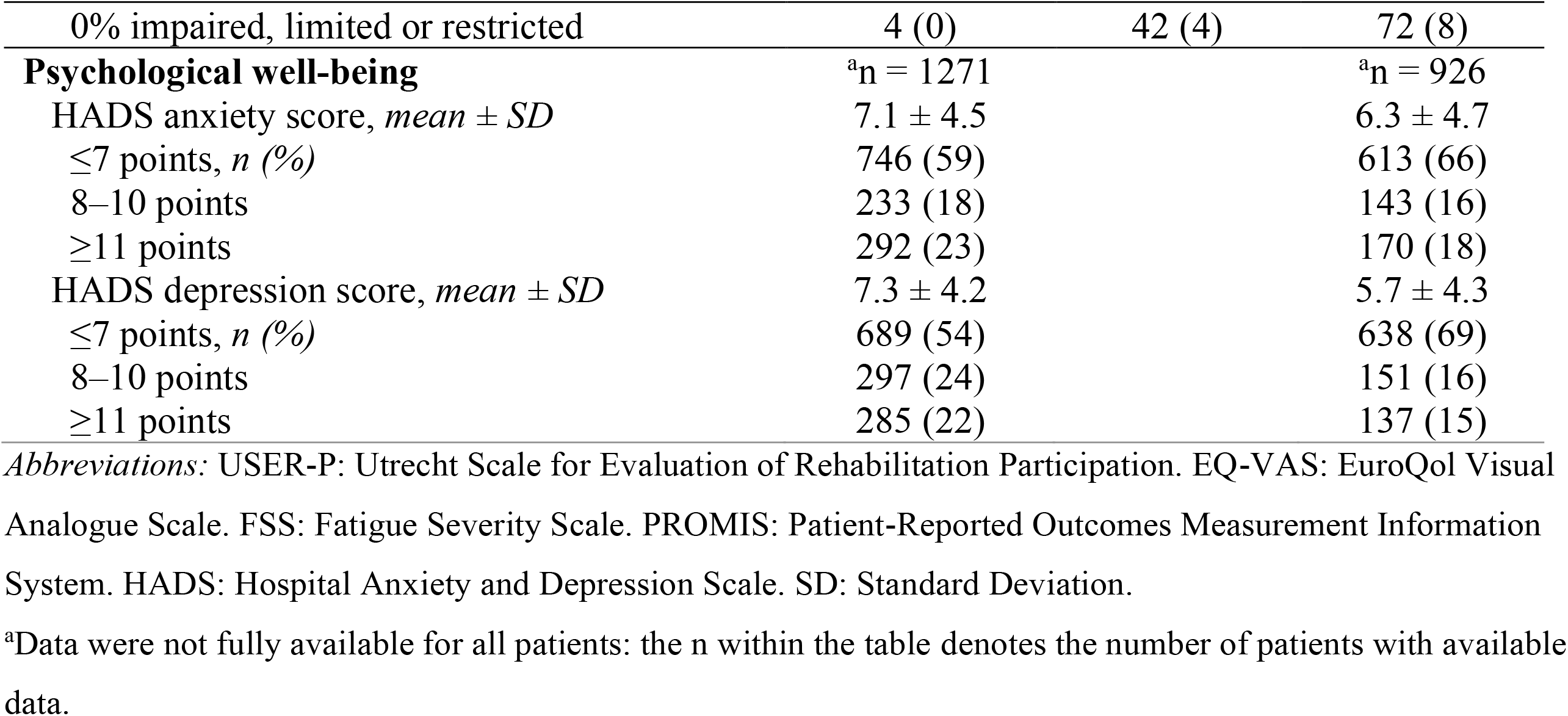
General outcome measures at the baseline, and after three and six months in patients recovering from COVID-19 receiving primary allied healthcare

### Patient-reported recovery over time

Table 3 shows the effect of time on the outcome measures. For all dependent variables, a random intercept model was the best-fitting model. No variables were significantly related to missing values in the outcome measures at any point in time. A significant effect of time was observed for all outcome measures at the three- and six-month follow-ups (*p* < 0.001). For participation, estimated mean differences of at least 2.9 points (*p* < 0.001) were observed for all three scales at all time points. For HRQoL, the mean increase was 9.0 points (95% CI 7.8 to 10.2) at three months and 12.3 points (95% CI 11.1 to 13.6) after six months. Furthermore, significant improvements were found for fatigue and physical functioning at all time points. The greatest improvements were seen after just three months for all outcome measures measured at both three and six months. Paired-samples t-tests showed mean differences at six months of –0.8 (95% CI –1.0 to –0.5) on the HADS anxiety score and –1.5 (95% CI –1.8 to – 1.3) on the HADS depression score.

**Table 3.**
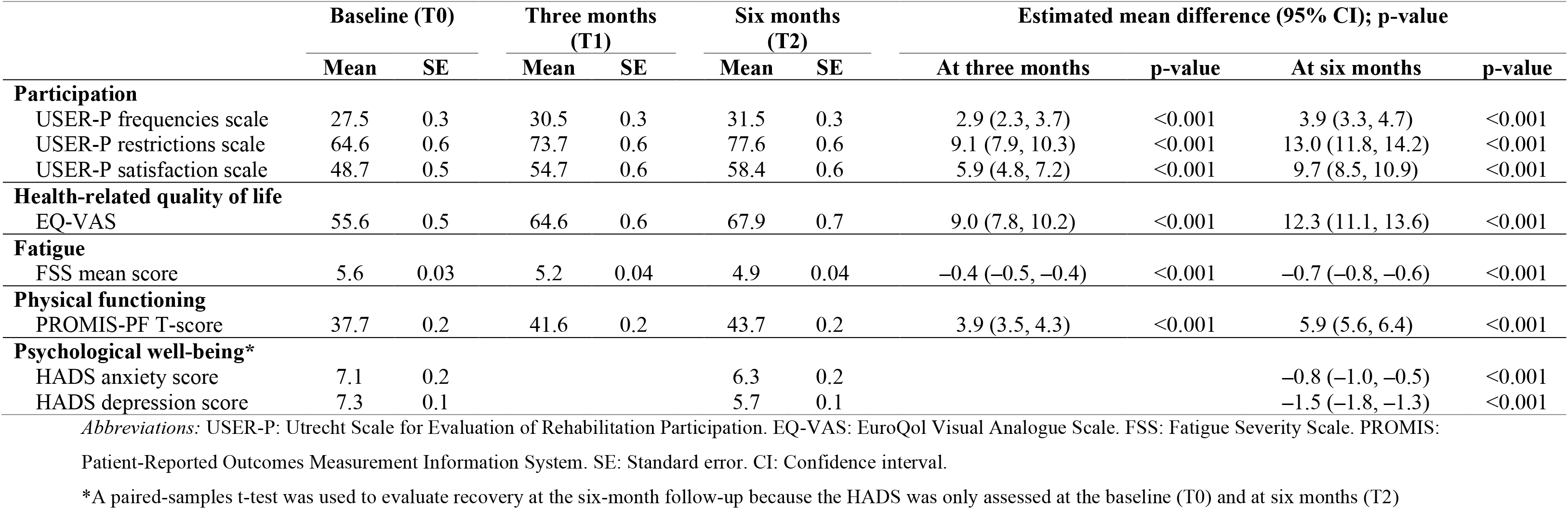
Results of linear mixed model analysis for the outcome measures participation, health-related quality of life, fatigue and physical functioning.

### Factors associated with the change in score in the main outcome measures

Multivariable regression models were estimated to identify factors associated with the change in scores between the baseline and the six-month follow-up in each outcome measure. Table 4 provides an overview of the final regression models. All univariable and multivariable regression models are shown in Supplemental Files 3 and 4. Having a worse baseline score was related to greater improvements for all outcome measures. For all three scales of the USER-P and physical functioning, patients admitted to hospital during the infection period of COVID-19 showed greater improvements in scores than non-hospitalised patients, even when correcting for the baseline scores. In terms of HRQoL, patients admitted to a hospital ward showed greater improvements than patients who had not been hospitalised, although no associations were found with ICU admission. Male participants showed greater improvements than female participants on all outcome measures, except for psychological well-being, for which no association was found for sex. The baseline age, BMI, comorbidities and smoking status were not significantly associated with the mean change in any of the outcome measures in our patient population.

**Table 4.** Multivariable linear regression models on the outcome measures.

| Outcome measure / Factor | $\beta$ | 95% confidence interval | p-value |
| --- | --- | --- | --- |
| **Participation (USER-P frequencies scale)** | **R<sup>2</sup> overall model: 0.272 ($p < 0.001$)** |  |  |
| Hospital admission |  |  | .001 |
| No | ref |  |  |
| Hospital ward | 2.556 | 0.851 – 4.262 | .003 |
| ICU | 3.079 | 0.611 – 5.547 | .015 |
| Baseline score | –0.496 | –0.558 – –0.435 | .000 |
| **Participation (USER-P restrictions scale)** | **R<sup>2</sup> overall model: 0.277 ($p < 0.001$)** |  |  |
| Sex |  |  | <0.001 |
| Male | ref |  |  |
| Female | –5.337 | –7.813 – –2.861 | <0.001 |
| Hospital admission |  |  | <0.001 |
| No | ref |  |  |
| Hospital ward | 3.581 | 0.316 – 6.845 | .032 |
| ICU | 9.165 | 4.522 – 13.809 | <0.001 |
| Baseline score | –0.462 | –0.520 – –0.405 | <0.001 |
| **Participation (USER-P satisfaction scale)** | **R<sup>2</sup> overall model: 0.159 ($p < 0.001$)** |  |  |
| Hospital admission |  |  | .003 |
| No | ref |  |  |
| Hospital ward | 3.577 | 0.356 – 6.798 | .030 |
| ICU | 6.728 | 2.144 – 11.311 | .004 |
| Baseline score | –0.402 | –0.467 – –0.338 | <0.001 |
| **Health-related quality of life (EQ-VAS)** | **R<sup>2</sup> overall model: 0.245 ($p < 0.001$)** |  |  |
| Sex |  |  | <0.001 |
| Male | ref |  |  |
| Female | –4.855 | –7.378 – –2.333 | <0.001 |
| Hospital admission |  |  | .097 |
| No | ref |  |  |
| Hospital ward | 3.594 | 0.231 – 6.957 | .036 |
| ICU | 2.106 | –2.615 – 6.827 | .381 |
| Baseline score | –0.524 | –0.589 – –0.459 | <0.001 |
| **Fatigue (FSS mean score)** | **R<sup>2</sup> overall model: 0.070 ($p < 0.001$)** |  |  |
| Sex |  |  | <0.001 |
| Male | ref |  |  |
| Female | 0.284 | 0.130 – 0.438 | <0.001 |
| Baseline score | –0.301 | –0.381 – –0.222 | <0.001 |
| **Physical functioning (PROMIS-PF T-score)** | **R<sup>2</sup> overall model: 0.064 ($p < 0.001$)** |  |  |
| Sex |  |  | <0.001 |
| Male | ref |  |  |
| Female | –2.342 | –3.341 – –1.343 | <0.001 |
| Hospital admission |  |  |  |
| No | ref |  | .004 |
| Hospital ward | 1.149 | –0.165 – 2.463 | .087 |
| ICU | 2.917 | 1.064 – 4.771 | .002 |
| Baseline score | –0.125 | –0.203 – –0.046 | .002 |
| **Psychological well-being (HADS anxiety)** | **R<sup>2</sup> overall model: 0.160 ($p < 0.001$)** |  |  |
| Baseline score | –0.354 | –0.407 – –0.301 | <0.001 |
| **Psychological well-being (HADS depression)** | **R<sup>2</sup> overall model: 0.179 ($p < 0.001$)** |  |  |
| Baseline score | –0.392 | –0.447 – –0.337 | <0.001 |

Abbreviations: USER-P: Utrecht Scale for Evaluation of Rehabilitation Participation. EQ-VAS: EuroQol Visual Analogue Scale. FSS: Fatigue Severity Scale. PROMIS: Patient-Reported Outcomes Measurement Information System. SE: Standard error. CI: Confidence interval. Ref: Reference value.

## Discussion

This study presents the first results of our evaluation of the recovery of our unique cohort of patients with COVID-19 receiving primary allied healthcare until their six-month follow-up. We explored which baseline characteristics were associated with the change in scores for the main outcome measures over this six-month period. Most patients showed a clinically relevant improvement in all outcome measures; however, despite improvement, many patients still experienced persistent problems in their daily lives, with limitations in physical and mental functioning. Having a worse baseline score, hospital admission and, for some outcome measures, male sex were associated with greater improvement between the baseline and the six-month follow-up; however, age, BMI, comorbidities and smoking status were not associated with the mean change in any of the outcome measures.

### Comparison with other studies

The majority of our patient population showed a clinically relevant improvement six months after starting treatment provided by one or more primary care allied health professional(s); nevertheless, a large group of patients experienced persistent problems in their daily lives. The mean EQ-VAS score of our patient population (67 points) remained well below the population norm in the Netherlands, which is 82 points.[41] These results are consistent with previous findings that HRQoL was impaired in the majority of post-COVID-19 patients.[12, 15, 17, 42-45] Persistent fatigue was highly prevalent among the patients included in our study, with 80% still reporting severe fatigue (measured with the FSS) after six months. These results are consistent with previous studies in patients recovering from COVID-19 showing that fatigue was the most common complaint,[5, 10, 14, 44, 46-48] even after six months.[13, 15, 49-51] The mean PROMIS T-score of our population (43.5 (SD 8.5)) remained well below the population norm in the Netherlands (mean score of 50 (SD 10)). These results are also consistent with previous studies,[15, 44] and indicate that persistent symptoms due to COVID-19 may lead to experienced limitations in physical functioning.

Relative to other outcome measures, a smaller percentage of patients showed a clinically relevant improvement in psychological well-being. This was due to an observed ceiling effect, as 59% and 54% of patients showed no indication of an anxiety disorder or depression at the baseline, respectively. Data from these patients is still informative however, as they could also have deteriorated throughout the follow-up period. With scores indicating a probable clinical diagnosis of anxiety disorder or depression in 18% and 15% of patients, respectively, after six months, our findings are similar to those reported in previous studies, which showed prevalence rates ranging from 11% to 40%.[8, 17, 43, 45, 48, 51-54]

We found that male participants showed greater improvements than female participants in participation, HRQoL, fatigue and physical functioning. These results are consistent with previous studies showing that female participants experience more persistent symptoms after a COVID-19 infection.[10, 14, 21, 42, 46, 48] Furthermore, patients admitted to the hospital for COVID-19 showed greater improvements than non-hospitalised patients in terms of participation, HRQoL and physical functioning, which is in line with previous studies.[12, 21, 42, 48] We observed no associations between fatigue and hospital admission, age, BMI, comorbidities or smoking status, which is also consistent with other studies,[10, 14, 48, 50, 51] indicating that fatigue is highly prevalent in patients recovering from COVID-19, irrespective of the severity of initial infection and patient characteristics. We found that having a worse baseline score was related to greater improvement in anxiety and depressive symptoms; however, no associations with any patient characteristics were found. Similar to our results, previous studies found no associations between the frequency of anxiety or depressive symptoms and disease severity or hospital admission.[10, 43, 45, 46, 54] In contrast, other studies found female sex [51, 55, 56] and older age [51, 53] to be predictors of anxiety or depressive symptoms in patients with COVID-19.

### Strengths and limitations

This is a unique comprehensive study presenting longitudinal results for patients recovering from a COVID-19 infection treated by one or more primary care allied health professionals. This large nationwide study used patient-reported outcome measures (PROMs) to evaluate the primary care allied health recovery trajectories, whereas previous studies mainly reported the prevalence of persistent symptoms in patients recovering with COVID-19. The use of PROMs can measure the quality of care based on the perceived health and functioning of patients, and leads to better communication and decision-making between health professionals and patients.[57-59]

Due to ethical considerations, this study did not include a control group to determine the potential effects of primary allied healthcare by comparing outcome measures with patients who did not receive this type of care. In addition, with a lack of available pre-COVID data for our population, it is difficult to draw conclusions about the impact of pre-existing conditions versus problems in the daily activities and participation of these patients due to their COVID-19 infection.

For the interpretation of the results, it is important to consider that the baseline measurement in this study was taken at the start of the treatment by one or more primary care allied health professional(s). It is possible that a patient had already experienced symptoms for some time and only consulted an allied health professional at a later stage. Additionally, it should be taken into account that not all patients received treatment by one or more allied health professional(s) during the entire six-month follow-up period of this study. Some patients received short-term treatment, while others were still receiving treatment at six months. Based on the inclusion period, which was between March and July 2021, our population most likely had the Wuhan or Alpha variant of the coronavirus SARS-CoV-2 [60]. Different variants may cause different symptoms, and the recovery trajectories of patients infected with other variants (e.g., Delta or Omicron) may differ from our population. A total of 25 patients dropped out during this study (Figure 1). Although a proportion of the patients did not complete all questionnaires, the response rates were still sufficient: 93% at the baseline, 68% after three months, and 67% after six months [61]. There was no selective missingness of data based on patient characteristics (including disease severity) and scores on the outcome measures (Supplemental File 1).

### Implications and future perspectives

The results of this study show that patients recovering from COVID-19 and receiving primary allied healthcare make progress in recovery, but many still experience limitations in their daily activities and participation after six months. The findings of our study provide reference values for healthcare providers and healthcare policy-makers about what to expect from the recovery of patients who receive or have received healthcare from one or more primary care allied health professional(s).

Future research and in-depth analyses of our data are needed to gain more insight into the outcome measures and recovery trajectories of patients recovering from COVID-19 who visit one or more primary care allied health professionals. Future papers will include the results after a 12-month follow-up, determining the related healthcare costs and the profession-specific outcomes per allied health discipline.

## Data Availability

According to international standards, data will be stored for 15 years. The data in this manuscript are anonymised and documented, and stored to be reusable after anonymisation. After publication of all studies of the ParaCOV consortium, the data can be reused upon reasonable request from the corresponding author.

## References

1. Taquet, M., et al., Incidence, co-occurrence, and evolution of long-COVID features: A 6-month retrospective cohort study of 273,618 survivors of COVID-19. PLoS medicine, 2021. 18(9): p. e1003773.

2. Webber, S.C., B.J. Tittlemier, and H.J. Loewen, Apparent discordance between the epidemiology of COVID-19 and recommended outcomes and treatments: a scoping review. Physical therapy, 2021. 101(11): p. pzab155.

3. Ballering, A.V., et al., Persistence of somatic symptoms after COVID-19 in the Netherlands: an observational cohort study. Lancet, 2022. 400(10350): p. 452–461.

4. Augustin, M., et al., Post-COVID syndrome in non-hospitalised patients with COVID-19: a longitudinal prospective cohort study. The Lancet Regional Health-Europe, 2021. 6: p. 100122.

5. Goërtz, Y.M., et al., Persistent symptoms 3 months after a SARS-CoV-2 infection: the post-COVID-19 syndrome? ERJ open research, 2020. 6(4).

6. NICE, COVID-19 rapid giudeline: managing the long-term effects of COVID-19. 2020, National Institute for Health and Care Excellence: Clinical Guidelines: London.

7. Shah, W., et al., Managing the long term effects of covid-19: summary of NICE, SIGN, and RCGP rapid guideline. bmj, 2021. 372.

8. Nalbandian, A., et al., Post-acute COVID-19 syndrome. Nature medicine, 2021. 27(4): p. 601–615.

9. Carfì, A., R. Bernabei, and F. Landi, Persistent symptoms in patients after acute COVID-19. Jama, 2020. 324(6): p. 603–605.

10. Sisó-Almirall, A., et al., Long Covid-19: proposed primary care clinical guidelines for diagnosis and disease management. International journal of environmental research and public health, 2021. 18(8): p. 4350.

11. Maxwell, E., Living with Covid19. National Institute for Health Research, 2020.

12. Malik, P., et al., Post-acute COVID-19 syndrome (PCS) and health-related quality of life (HRQoL)—A systematic review and meta-analysis. Journal of medical virology, 2022. 94(1): p. 253–262.

13. Van Herck, M., et al., Severe Fatigue in Long COVID: Web-Based Quantitative Follow-up Study in Members of Online Long COVID Support Groups. Journal of Medical Internet Research, 2021. 23(9): p. e30274.

14. Townsend, L., et al., Persistent fatigue following SARS-CoV-2 infection is common and independent of severity of initial infection. Plos one, 2020. 15(11): p. e0240784.

15. Tabacof, L., et al., Post-acute COVID-19 Syndrome Negatively Impacts Physical Function, Cognitive Function, Health-Related Quality of Life, and Participation. American journal of physical medicine & rehabilitation, 2022. 101(1): p. 48.

16. Humphreys, H., et al., Long COVID and the role of physical activity: a qualitative study. BMJ open, 2021. 11(3): p. e047632.

17. Huang, C., et al., 6-month consequences of COVID-19 in patients discharged from hospital: a cohort study. The Lancet, 2021. 397(10270): p. 220–232.

18. Nguyen, H.C., et al., People with suspected COVID-19 symptoms were more likely depressed and had lower health-related quality of life: the potential benefit of health literacy. Journal of clinical medicine, 2020. 9(4): p. 965.

19. Zhang, Y. and Z.F. Ma, Impact of the COVID-19 pandemic on mental health and quality of life among local residents in Liaoning Province, China: A cross-sectional study. International journal of environmental research and public health, 2020. 17(7): p. 2381.

20. Qu, G., et al., Health-related quality of life of COVID-19 patients after discharge: a multicenter follow-up study. Journal of clinical nursing, 2021. 30(11-12): p. 1742–1750.

21. Arab-Zozani, M., et al., Health-related quality of life and its associated factors in COVID-19 patients. Osong public health and research perspectives, 2020. 11(5): p. 296.

22. KNGF-Standpunt Fysiotherapie bij patienten met COVID-19: Royal Duth Society for Physical Therapy (KNGF). 2020; Available from: https://www.kngf.nl/binaries/content/assets/kennisplatform/onbeveiligd/coronavirus/kngf_standpunt_fysiotherapie_covid-19_v3.0_22022022.pdf.

23. Position statment COVID-19 version 1.4: Dutch Society for speech-language therapy (NVLF). 2020; Available from: https://www.nvlf.nl/wp-content/uploads/sites/2/2020/06/Factsheet-Intramurale-instellingen.pdf.

24. Handreiking ergotherapie bij COVID-19 cliënten in de herstelfase Dutch society for occupational therapy (EN). 2021; Available from: https://info.ergotherapie.nl/file/download/default/6A5E0AC0401E6972DA637BB919F13500/26-01-21%20-%20Handreiking%20ergotherapie%20bij%20COVID-19%20in%20de%20herstelfase%20-%20versie%20januari%202021.pdf.

25. Treatment plan of dietitian at COVID-19 after hospital discharge The recovery phase after discharge from hospital: dietetics treatment in (Post) COVID-19 patients in a rehabilitation center: Dutch society for dietitians (NVD). 2021; Available from: https://nvdietist.nl/artikelen/behandelplan-van-dietist-binnen-paramedische-herstelzorg-covid-19/.

26. De Bie, R.A., et al., Evaluation of Allied Healthcare in Patients Recovering from Covid-19: Study Protocol and Baseline Data of a National Prospective Cohort Study. Journal of Rehabilitation Medicine, 2022: p. jrm00309-jrm00309.

27. CDC. Interim Clinical Guidance for Management of Patients with Confirmed Coronavirus Disease (COVID-19). 2022; Available from: https://www.cdc.gov/coronavirus/2019-ncov/hcp/clinical-guidance-management-patients.html.

28. WHO. Body Mass Index - BMI. Available from: https://www.who.int/europe/news-room/fact-sheets/item/a-healthy-lifestyle---who-recommendations.

29. Post, M.W., et al., Validity of the utrecht scale for evaluation of rehabilitation-participation. Disability and rehabilitation, 2012. 34(6): p. 478–485.

30. Herdman, M., et al., Development and preliminary testing of the new five-level version of EQ-5D (EQ-5D-5L). Quality of life research, 2011. 20(10): p. 1727–1736.

31. Zanini, A., et al., Estimation of minimal clinically important difference in EQ-5D visual analog scale score after pulmonary rehabilitation in subjects with COPD. Respiratory care, 2015. 60(1): p. 88–95.

32. Valko, P.O., et al., Validation of the fatigue severity scale in a Swiss cohort. Sleep, 2008. 31(11): p. 1601–1607.

33. Rooney, S., et al., Minimally important difference of the fatigue severity scale and modified fatigue impact scale in people with multiple sclerosis. Multiple sclerosis and related disorders, 2019. 35: p. 158–163.

34. Cella, D., et al., The Patient-Reported Outcomes Measurement Information System (PROMIS): progress of an NIH Roadmap cooperative group during its first two years. Medical care, 2007. 45(5 Suppl 1): p. S3.

35. Terwee, C., et al., Dutch–Flemish translation of 17 item banks from the patient-reported outcomes measurement information system (PROMIS). Quality of Life Research, 2014. 23(6): p. 1733–1741.

36. Sandvall, B., et al., Minimal clinically important difference for PROMIS physical function in patients with distal radius fractures. The Journal of Hand Surgery, 2019. 44(6): p. 454-459. e1.

37. Snaith, R.P., The hospital anxiety and depression scale. Health and quality of life outcomes, 2003. 1(1): p. 1–4.

38. Zigmond, A.S. and R.P. Snaith, The hospital anxiety and depression scale. Acta psychiatrica scandinavica, 1983. 67(6): p. 361–370.

39. Lemay, K.R., et al., Establishing the minimal clinically important difference for the hospital anxiety and depression scale in patients with cardiovascular disease. Journal of cardiopulmonary rehabilitation and prevention, 2019. 39(6): p. E6–E11.

40. Steyerberg, E.W., Clinical Prediction Models: A Practical Approach to Development, Validation, and Updating. 2009, New York: Springer Sciences + Business Media, LLC.

41. Janssen, B. and A. Szende, Population norms for the EQ-5D. Self-reported population health: an international perspective based on EQ-5D, 2014: p. 19–30.

42. Garrigues, E., et al., Post-discharge persistent symptoms and health-related quality of life after hospitalization for COVID-19. Journal of Infection, 2020. 81(6): p. e4–e6.

43. Rass, V., et al., Neurological outcome and quality of life 3 months after COVID-19: A prospective observational cohort study. European journal of neurology, 2021. 28(10): p. 3348–3359.

44. Vaes, A.W., et al., Recovery from COVID-19: a sprint or marathon? 6-month follow-up data from online long COVID-19 support group members. ERJ open research, 2021. 7(2).

45. Van den Borst, B., et al., Comprehensive health assessment 3 months after recovery from acute coronavirus disease 2019 (COVID-19). Clinical Infectious Diseases, 2021. 73(5): p. e1089–e1098.

46. Ceban, F., et al., Fatigue and cognitive impairment in Post-COVID-19 Syndrome: A systematic review and meta-analysis. Brain, behavior, and immunity, 2022. 101: p. 93–135.

47. Pavli, A., M. Theodoridou, and H.C. Maltezou, Post-COVID syndrome: Incidence, clinical spectrum, and challenges for primary healthcare professionals. Archives of medical research, 2021. 52(6): p. 575–581.

48. Shanbehzadeh, S., et al., Physical and mental health complications post-COVID-19: Scoping review. Journal of psychosomatic research, 2021. 147: p. 110525.

49. Alkodaymi, M.S., et al., Prevalence of post-acute COVID-19 syndrome symptoms at different follow-up periods: A systematic review and meta-analysis. Clinical Microbiology and Infection, 2022.

50. Ezzat, M.M. and A.A. Elsherif, Prevalence of fatigue in patients post COVID-19. European Journal of Molecular & Clinical Medicine, 2021. 8(3): p. 1330–1340.

51. Menges, D., et al., Burden of post-COVID-19 syndrome and implications for healthcare service planning: A population-based cohort study. PloS one, 2021. 16(7): p. e0254523.

52. González, J., et al., Pulmonary function and radiologic features in survivors of critical COVID-19: a 3-month prospective cohort. Chest, 2021. 160(1): p. 187–198.

53. Morin, L., et al., Four-month clinical status of a cohort of patients after hospitalization for COVID-19. Jama, 2021. 325(15): p. 1525–1534.

54. Renaud-Charest, O., et al., Onset and frequency of depression in post-COVID-19 syndrome: A systematic review. Journal of psychiatric research, 2021. 144: p. 129–137.

55. Mazza, M.G., et al., Persistent psychopathology and neurocognitive impairment in COVID-19 survivors: effect of inflammatory biomarkers at three-month follow-up. Brain, behavior, and immunity, 2021. 94: p. 138–147.

56. Righi, E., et al., Determinants of persistence of symptoms and impact on physical and mental wellbeing in Long COVID: A prospective cohort study. Journal of Infection, 2022. 84(4): p. 566–572.

57. Kessel, P.v., M. Triemstra, and D.d. Boer, Handreiking voor het meten van kwaliteit van zorg met Patient Reported Outcome Measures. 2014.

58. Nelson, E.C., et al., Patient reported outcome measures in practice. Bmj, 2015. 350.

59. Gibbons, C., et al., Routine provision of feedback from patient-reported outcome measurements to healthcare providers and patients in clinical practice. Cochrane Database Syst Rev, 2021. 10(10): p. Cd011589.

60. RIVM. Coronadashboard - Varianten van het coronavirus. 2022; Available from: https://coronadashboard.rijksoverheid.nl/landelijk/varianten.

61. Nulty, D.D., The adequacy of response rates to online and paper surveys: what can be done? Assessment & evaluation in higher education, 2008. 33(3): p. 301–314.

