## Supplemental File for "Evaluation of primary allied healthcare in patients recovering from COVID-19: first results after six months follow-up in a Dutch nationwide prospective cohort study"

**Supplemental File 1**. Missingness at random on general outcome measures at baseline, three and six months in patients recovering from COVID-19 receiving primary allied healthcare

| | Total group | Lost-to-follow-up after T0 |
|---|---|---|
| **Participation**, *mean ± SD* | [a]n = 1271 | [a]n = 248 |
| USER-P frequencies scale | 27.5 ± 10.3 | 27.6 ± 10.8 |
| USER-P restrictions scale | 65.8 ± 20.1 | 64.8 ± 20.8 |
| USER-P satisfaction scale | 48.6 ± 17.8 | 49.3 ± 18.6 |
| **Health-related quality of life** | [a]n = 1289 | [a]n = 247 |
| EQ-VAS, *mean ± SD* | 55.5 ± 17.8 | 58.8 ± 17.4 |
| **Fatigue** | [a]n = 1281 | [a]n = 243 |
| FSS mean score, *mean ± SD* | 5.6 ± 1.0 | 5.4 ± 1.1 |
| ≥4 points, *n (%)* | 1205 (94) | 226 (93) |
| **Physical functioning** | [a]n = 1279 | [a]n = 245 |
| PROMIS-PF T-score, *mean ± SD* | 37.7 ± 6.0 | 38.0 ± 6.3 |
| 100% impaired, limited or restricted | 2 (0) | 1 (0) |
| 80-99% impaired, limited or restricted | 427 (33) | 73 (30) |
| 60-79% impaired, limited or restricted | 452 (35) | 84 (34) |
| 40-59% impaired, limited or restricted | 259 (20) | 61 (25) |
| 20-39% impaired, limited or restricted | 97 (8) | 16 (7) |
| 1-19% impaired, limited or restricted | 39 (3) | 9 (4) |
| 0% impaired, limited or restricted | 4 (0) | 1 (0) |
| **Psychological well-being** | [a]n = 1271 | [a]n = 370 |
| HADS anxiety score, *mean ± SD* | 7.1 ± 4.5 | 7.1 ± 4.5 |
| ≤7 points, *n (%)* | 746 (59) | 213 (58) |
| 8-10 points | 233 (18) | 68 (18) |
| ≥11 points | 292 (23) | 89 (24) |
| HADS depression score, *mean ± SD* | 7.3 ± 4.2 | 7.4 ± 4.2 |
| ≤7 points, *n (%)* | 689 (54) | 202 (55) |
| 8-10 points | 297 (24) | 89 (24) |
| ≥11 points | 285 (22) | 79 (21) |

*Abbreviations:* USER-P: Utrecht Scale for Evaluation of Rehabilitation Participation. EQ-VAS: EuroQol Visual Analogue Scale. FSS: Fatigue Severity Scale. PROMIS: Patient-Reported Outcomes Measurement Information System. HADS: Hospital Anxiety and Depression Scale. SD: Standard Deviation.

[a]Data were not fully available for all patients: the n within the table depicts the number of patients with available data.

**Supplemental File 2.** Clinically relevant improvement at six months follow-up of patients recovering from COVID-19 receiving primary allied healthcare.

| **Outcome measures** | **Baseline vs. six months follow-up**, *n (%)* |
|---|---|
| **USER-P restrictions scale** | [a]n = 890 |
| Clinically relevant improvement | 576 (65) |
| No clinically relevant improvement | 193 (22) |
| Clinically relevant deterioration | 121 (13) |
| **USER-P satisfaction scale** | [a]n = 891 |
| Clinically relevant improvement | 543 (61) |
| No clinically relevant improvement | 182 (20) |
| Clinically relevant deterioration | 166 (19) |
| **EQ-VAS** | [a]n = 908 |
| Clinically relevant improvement | 540 (60) |
| No clinically relevant improvement | 255 (28) |
| Clinically relevant deterioration | 113 (12) |
| **FSS mean score** | [a]n = 904 |
| Clinically relevant improvement | 490 (54) |
| No clinically relevant improvement | 319 (35) |
| Clinically relevant deterioration | 95 (11) |
| **PROMIS-PF T-score** | [a]n = 902 |
| Clinically relevant improvement | 517 (57) |
| No clinically relevant improvement | 357 (40) |
| Clinically relevant deterioration | 28 (3) |
| **HADS anxiety score** | [a]n = 901 |
| Clinically relevant improvement | 345 (38) |
| No clinically relevant improvement | 326 (36) |
| Clinically relevant deterioration | 230 (26) |
| **HADS depression score** | [a]n = 901 |
| Clinically relevant improvement | 428 (47) |
| No clinically relevant improvement | 303 (34) |
| Clinically relevant deterioration | 170 (19) |

*Abbreviations:* USER-P: Utrecht Scale for Evaluation of Rehabilitation Participation. EQ-VAS: EuroQol Visual Analogue Scale. FSS: Fatigue Severity Scale. PROMIS: Patient-Reported Outcomes Measurement Information System. HADS: Hospital Anxiety and Depression Scale.

[a]Data were not fully available for all patients: the n within the table depicts the number of patients with available data.

**Supplemental File 3**. Univariable logistic regression models on outcome measures.

### 1. USER-P

#### 1.1. Frequencies scale

| Factor | β | 95% Confidence Interval | p-value |
|---|---|---|---|
| Age | 0.086 | 0.027 – 0.144 | .004 |
| Sex | | | <0.001 |
| Male | ref | | |
| Female | -2.582 | -4.021 – -1.142 | <0.001 |
| Hospital admission | | | <0.001 |
| No | ref | | |
| Hospital ward | 4.453 | 2.522 – 6.384 | <0.001 |
| ICU | 7.034 | 4.270 – 9.798 | <0.001 |
| BMI | | | .818 |
| Normal/underweight | ref | | |
| Overweight | 0.612 | -1.290 – 2.515 | .528 |
| Obese | 0.382 | -1.601 – 2.365 | .705 |
| Comorbidities | | | .765 |
| 0 | ref | | |
| 1 | -0.417 | -2.013 – 1.070 | .548 |
| ≥2 | 0.303 | -2.004 – 2.610 | .797 |
| Smoking status | | | .861 |
| Never | ref | | |
| Former | -0.092 | -2.299 – 2.115 | .935 |
| Current | -0.957 | -4.402 – 2.487 | .586 |
| Baseline score | -0.514 | -0.572 – -0.455 | <0.001 |

#### 1.2. Restrictions scale

| Factor | β | 95% Confidence Interval | p-value |
|---|---|---|---|
| Age | -0.008 | -0.118 – 0.102 | .891 |
| Sex | | | .001 |
| Male | ref | | |
| Female | -4.403 | -7.099 – -1.707 | .001 |
| Hospital admission | | | <0.001 |
| No | ref | | |
| Hospital ward | 5.442 | 1.851 – 9.033 | .003 |
| ICU | 15.730 | 10.589 – 20.870 | <0.001 |
| BMI | | | .463 |
| Normal/underweight | ref | | |
| Overweight | 1.783 | -1.528 – 5.473 | .269 |
| Obese | 1.974 | -1.674 – 5.622 | .288 |

| | | | |
|---|---|---|---|
| Comorbidities | | | .393 |
| 0 | ref | | |
| 1 | -1.798 | -4.676 – 1.080 | .220 |
| ≥2 | -1.952 | -6.258 – 2.354 | .374 |
| Smoking status | | | .703 |
| Never | ref | | |
| Former | 1.628 | -2.481 – 5.736 | .437 |
| Current | -0.829 | -7.242 – 5.584 | .800 |
| Baseline score | -0.454 | -0.509 – -0.399 | <0.001 |

#### 1.3. Satisfaction scale

| Factor | β | 95% Confidence Interval | p-value |
|---|---|---|---|
| Age | 0.097 | -0.201 – 0.006 | .066 |
| Sex | | | .235 |
| Male | ref | | |
| Female | -1.547 | -4.101 – 1.007 | .235 |
| Hospital admission | | | .122 |
| No | ref | | |
| Hospital ward | 1.120 | -2.354 – 4.594 | .527 |
| ICU | 5.099 | 0.126 – 10.072 | .045 |
| BMI | | | .986 |
| Normal/underweight | ref | | |
| Overweight | -0.289 | -3.688 – 3.111 | .868 |
| Obese | -0.121 | -3.664 – 3.421 | .946 |
| Comorbidities | | | .447 |
| 0 | ref | | |
| 1 | -1.756 | -4.470 – 0.958 | .204 |
| ≥2 | -0.608 | -4.669 – 3.453 | .769 |
| Smoking status | | | .440 |
| Never | ref | | |
| Former | 2.185 | -1.714 – 6.083 | .272 |
| Current | 2.306 | -3.779 – 8.391 | .457 |
| Baseline score | -0.387 | -0.449 – -0.326 | <0.001 |

### 2. EQ-VAS

| Factor | β | 95% Confidence Interval | p-value |
|---|---|---|---|
| Age | -0.014 | -0.123 – 0.096 | .809 |
| Sex | | | .003 |
| Male | ref | | |
| Female | -4.051 | -6.756 – -1.346 | .003 |
| Hospital admission | | | .093 |
| No | ref | | |

| | | | |
|---|---|---|---|
| Hospital ward | 3.944 | 0.244 – 7.644 | .037 |
| ICU | 2.300 | -2.974 – 7.575 | .392 |
| BMI | | | .814 |
| Normal/underweight | ref | | |
| Overweight | -0.556 | -4.092 – 2.980 | .758 |
| Obese | 0.592 | -3.091 – 4.274 | .753 |
| Comorbidities | | | .477 |
| 0 | ref | | |
| 1 | -0.132 | -3.017 – 2.752 | .928 |
| ≥2 | -2.624 | -6.921 – 1.674 | .231 |
| Smoking status | | | .185 |
| Never | ref | | |
| Former | 3.101 | -0.993 – 7.196 | .137 |
| Current | -3.163 | -9.535 – 3.264 | .336 |
| Baseline score | -0.502 | -0.563 – -0.440 | <0.001 |

#### 3. FSS mean score

| Factor | β | 95% Confidence Interval | p-value |
|---|---|---|---|
| Age | -0.004 | -0.010 – 0.003 | .269 |
| Sex | | | .014 |
| Male | ref | | |
| Female | 0.198 | 0.040 – 0.355 | .014 |
| Hospital admission | | | .067 |
| No | ref | | |
| Hospital ward | -0.224 | -0.437 – -0.011 | .039 |
| ICU | 0.131 | -0.174 – 0.435 | .401 |
| BMI | | | .621 |
| Normal/underweight | ref | | |
| Overweight | -0.035 | -0.244 – 0.175 | .746 |
| Obese | -0.106 | -0.324 – 0.112 | .341 |
| Comorbidities | | | .736 |
| 0 | ref | | |
| 1 | -0.018 | -0.186 – 0.149 | .829 |
| ≥2 | 0.086 | -0.163 – 0.335 | .499 |
| Smoking status | | | .466 |
| Never | ref | | |
| Former | 0.011 | -0.225 – 0.248 | .925 |
| Current | -0.234 | -0.609 – 0.142 | .223 |
| Baseline score | -0.313 | -0.390 – -0.237 | <0.001 |

#### 4. PROMIS-PF T-score

| Factor | β | 95% Confidence Interval | p-value |
|---|---|---|---|
| Age | 0.007 | -0.031 – 0.045 | .728 |
| Sex | | | <0.001 |
| Male | ref | | |
| Female | -2.612 | -3.542 – -1.682 | <0.001 |
| Hospital admission | | | <0.001 |
| No | ref | | |
| Hospital ward | 2.129 | 0.856 – 3.401 | .001 |
| ICU | 4.242 | 2.435 – 6.050 | <0.001 |
| BMI | | | .709 |
| Normal/underweight | ref | | |
| Overweight | .496 | -0.771 – 1.764 | .442 |
| Obese | .449 | -0.872 – 1.770 | .505 |
| Comorbidities | | | .771 |
| 0 | ref | | |
| 1 | -0.119 | -1.125 – 0.886 | .816 |
| ≥2 | -0.551 | -2.053 – 0.951 | .472 |

| | | | |
|---|---|---|---|
| Smoking status | | | .361 |
| Never | ref | | |
| Former | 1.039 | -0.400 – 2.479 | .157 |
| Current | -0.074 | -2.322 – 2.174 | .948 |
| Baseline score | -0.108 | -0.181 – -0.034 | .004 |

### 5. HADS

#### 5.1. Anxiety score

| Factor | β | 95% Confidence Interval | p-value |
|---|---|---|---|
| Age | -0.009 | -0.032 – 0.014 | .429 |
| Sex | | | .796 |
| Male | ref | | |
| Female | 0.074 | -0.486 – 0.634 | .796 |
| Hospital admission | | | .103 |
| No | ref | | |
| Hospital ward | 0.173 | -0.590 – 0.936 | .656 |
| ICU | 1.167 | 0.089 – 2.245 | .034 |
| BMI | | | .860 |
| Normal/underweight | ref | | |
| Overweight | 0.007 | -0.710 – 0.724 | .985 |
| Obese | -0.172 | -0.917 – 0.573 | .650 |
| Comorbidities | | | .773 |
| 0 | ref | | |
| 1 | -0.208 | -0.804 – 0.389 | .495 |
| ≥2 | -0.172 | -1.047 – 0.703 | .699 |
| Smoking status | | | .070 |
| Never | ref | | |
| Former | 0.104 | -0.732 – 0.941 | .806 |
| Current | -1.584 | -2.951 – -0.216 | .023 |
| Baseline score | -0.354 | -0.407 – -0.301 | <0.001 |

#### 5.2. Depression score

| Factor | β | 95% Confidence Interval | p-value |
|---|---|---|---|
| Age | 0.014 | -0.008 – 0.036 | .221 |
| Sex | | | .430 |
| Male | ref | | |
| Female | 0.219 | -0.327 – 0.765 | .430 |
| Hospital admission | | | .033 |
| No | ref | | |
| Hospital ward | 0.408 | -0.335 – 1.150 | .282 |
| ICU | 1.335 | 0.285 – 2.385 | .013 |
| BMI | | | .779 |
| Normal/underweight | ref | | |
| Overweight | 0.176 | -0.552 – 0.904 | .635 |
| Obese | 0.268 | -0.488 – 1.024 | .487 |
| Comorbidities | | | .431 |
| 0 | ref | | |
| 1 | -0.166 | -0.748 – 0.416 | .576 |

| | | | |
|---|---|---|---|
| ≥2 | 0.432 | -0.421 – 1.285 | .321 |
| Smoking status | | | .084 |
| Never | ref | | |
| Former | -0.072 | -0.888 – 0.754 | .863 |
| Current | -1.516 | -2.851 – -0.181 | .026 |
| Baseline score | -0.392 | -0.447 – -0.337 | <0.001 |

**Supplemental File 4**. Multivariable logistic regression models on outcome measures.

### 1. USER-P

#### 1.1. Frequencies scale

| Factor | β | 95% Confidence Interval | p-value |
|---|---|---|---|
| Age | -0.057 | -0.112 – -0.001 | .045 |
| Sex | | | .129 |
| Male | ref | | |
| Female | -1.043 | -2.392 – 0.306 | .129 |
| Hospital admission | | | .003 |
| No | ref | | |
| Hospital ward | 2.704 | 0.898 – 4.511 | .003 |
| ICU | 2.973 | 0.457 – 5.489 | .021 |
| Baseline score | -0.506 | -0.568 – -0.443 | <0.001 |

$R^2$ overall model: 0.277 ($p < 0.001$)

| Factor | β | 95% Confidence Interval | p-value |
|---|---|---|---|
| Age | -0.045 | -0.099 – 0.009 | .103 |
| Hospital admission | | | .001 |
| No | ref | | |
| Hospital ward | 2.951 | 1.182 – 4.720 | .001 |
| ICU | 3.285 | 0.807 – 5.763 | .009 |
| Baseline score | -0.506 | -0.569 – -0.444 | <0.001 |

$R^2$ overall model: 0.275 ($p < 0.001$)

| Factor | β | 95% Confidence Interval | p-value |
|---|---|---|---|
| Hospital admission | | | .001 |
| No | ref | | |
| Hospital ward | 2.556 | 0.851 – 4.262 | .003 |
| ICU | 3.079 | 0.611 – 5.547 | .015 |
| Baseline score | -0.496 | -0.558 – -0.435 | <0.001 |

$R^2$ overall model: 0.272 ($p < 0.001$)

#### 1.2. Restrictions scale

| Factor | β | 95% Confidence Interval | p-value |
|---|---|---|---|
| Sex | | | <0.001 |
| Male | ref | | |
| Female | -5.337 | -7.813 – -2.861 | <0.001 |
| Hospital admission | | | <0.001 |
| No | ref | | |
| Hospital ward | 3.581 | 0.316 – 6.845 | .032 |
| ICU | 9.165 | 4.522 – 13.809 | <0.001 |

| | | | |
|---|---|---|---|
| Baseline score | -0.462 | -0.520 – -0.405 | <0.001 |

$R^2$ overall model: 0.277 ($p < 0.001$)

1.3. Satisfaction scale

| Factor | β | 95% Confidence Interval | p-value |
|---|---|---|---|
| Age | 0.005 | -0.099 – 0.108 | .930 |
| Hospital admission | | | .005 |
| No | ref | | |
| Hospital ward | 3.536 | 0.189 – 6.884 | .038 |
| ICU | 6.701 | 2.076 – 11.326 | .005 |
| Baseline score | -0.403 | -0.470 – -0.337 | <0.001 |

$R^2$ overall model: 0.159 ($p < 0.001$)

| Factor | β | 95% Confidence Interval | p-value |
|---|---|---|---|
| Hospital admission | | | .003 |
| No | ref | | |
| Hospital ward | 3.577 | 0.356 – 6.798 | .030 |
| ICU | 6.728 | 2.144 – 11.311 | .004 |
| Baseline score | -0.402 | -0.467 – -0.338 | <0.001 |

$R^2$ overall model: 0.159 ($p < 0.001$)

### 2. EQ-VAS

| Factor | β | 95% Confidence Interval | p-value |
|---|---|---|---|
| Sex | | | <0.001 |
| Male | ref | | |
| Female | -4.855 | -7.378 – -2.333 | <0.001 |
| Hospital admission | | | .097 |
| No | ref | | |
| Hospital ward | 3.594 | 0.231 – 6.957 | .036 |
| ICU | 2.106 | -2.615 – 6.827 | .381 |
| Baseline score | -0.524 | -0.589 – -0.459 | <0.001 |

$R^2$ overall model: 0.245 ($p < 001$)

### 3. FSS mean score

| Factor | β | 95% Confidence Interval | p-value |
|---|---|---|---|
| Sex | | | .002 |
| Male | ref | | |
| Female | 0.255 | 0.093 – 0.418 | .002 |
| Hospital admission | | | .145 |
| No | ref | | |
| Hospital ward | -0.208 | -0.423 – 0.007 | .058 |
| ICU | 0.029 | -0.277 – 0.335 | .852 |
| Baseline score | -0.303 | -0.383 – -0.223 | <0.001 |

$R^2$ overall model: 0.075 ($p < 001$)

| Factor | β | 95% Confidence Interval | p-value |
| --- | --- | --- | --- |
| Sex | | | <0.001 |
| Male | ref | | |
| Female | 0.284 | 0.130 – 0.438 | <0.001 |
| Baseline score | -0.301 | -0.381 – -0.222 | <0.001 |

$R^2$ overall model: 0.070 ($p$ < 001)

### 4. PROMIS-PF T-score

| Factor | β | 95% Confidence Interval | p-value |
|---|---|---|---|
| Sex | | | <0.001 |
| Male | ref | | |
| Female | -2.342 | -3.341 – -1.343 | <0.001 |
| Hospital admission | | | |
| No | ref | | .004 |
| Hospital ward | 1.149 | -0.165 – 2.463 | .087 |
| ICU | 2.917 | 1.064 – 4.771 | .002 |
| Baseline score | -0.125 | -0.203 – -0.046 | .002 |

$R^2$ overall model: 0.064 ($p$ < 001)

### 1. HADS

#### 1.1. Anxiety score

| Factor | β | 95% Confidence Interval | p-value |
|---|---|---|---|
| Hospital admission | | | .154 |
| No | ref | | |
| Hospital ward | -0.320 | -1.026 – 0.387 | .375 |
| ICU | 0.810 | -0.184 – 1.804 | .110 |
| Baseline score | -0.346 | -0.402 – -0.291 | <0.001 |

$R^2$ overall model: 0.158 ($p$ < 001)

| Factor | β | 95% Confidence Interval | p-value |
|---|---|---|---|
| Baseline score | -0.354 | -0.407 – -0.301 | <0.001 |

$R^2$ overall model: 0.160 ($p$ < 001)

#### 1.2. Depression score

| Factor | β | 95% Confidence Interval | p-value |
|---|---|---|---|
| Hospital admission | | | .317 |
| No | ref | | |
| Hospital ward | -0.007 | -0.686 – 0.672 | .984 |
| ICU | 0.735 | -0.225 – 1.695 | .133 |
| Baseline score | -0.386 | -0.444 – -0.328 | <0.001 |

$R^2$ overall model: 0.179 ($p$ < 001)

| Factor | β | 95% Confidence Interval | p-value |
|---|---|---|---|
| Baseline score | -0.392 | -0.447 – -0.337 | <0.001 |

$R^2$ overall model: 0.179 ($p$ < 001)
